# Extracellular Vesicle-Associated Syndecan-1 Differentiates Pediatric Brain Tumor Patients with High-Grade from Low-Grade Pilocytic Astrocytoma

**DOI:** 10.64898/2026.01.22.26344286

**Authors:** Jonas Klejs Hemmingsen, Josephine Elgaard Johansen, Mads Zippor, Bradley Whitehead, Anders Toftegaard Boysen, Hannah Weissinger, Mette Galsgaard Malle, Kenneth A. Howard, Srinivas Gopala, Peter Nejsum, Torben Stamm Mikkelsen, Vineesh Indira Chandran

## Abstract

Reliable non-invasive biomarkers for tumor grading and disease monitoring in pediatric brain tumors are an unmet clinical need. Circulating extracellular vesicles (EVs) carrying molecular cargo reflective of tumor biology, offer promise as liquid biopsy tools. We have previously discovered EV-associated Syndecan-1 (SDC1) to be overexpressed in malignant brain tumors, but its value as a biomarker in pediatric disease remains unclear. In this study, plasma EVs were isolated from pediatric brain tumor patients (n=60) by size-exclusion chromatography and characterized using cryo-electron microscopy, nanoflow cytometry, immunoblotting, and single-vesicle total internal reflection fluorescence imaging. EV-associated SDC1 (EV-SDC1) was quantified and analyzed in relation to tumor grade, subtype, surgical resection status, and tumor volume. EV-SDC1 levels were significantly elevated in high-grade (ependymoma, diffuse midline glioma, and atypical teratoid/rhabdoid tumor (AT/RT)) compared with low-grade pilocytic astrocytoma tumors and robustly discriminated grade 3 tumors from pilocytic astrocytoma (AUROC 1.00). Independent validation using transcriptomic data from the Open Pediatric Brain Tumor Atlas showed SDC1 mRNA levels to effectively distinguish high grade (ependymoma, medulloblastoma, diffuse midline glioma, and AT/RT) from pilocytic astrocytoma patients. Furthermore, EV-SDC1 levels decreased following complete tumor resection but remained elevated in patients with residual disease or recurrence. Collectively, circulating SDC1-positive EVs represents a clinically informative biomarker reflecting tumor aggressiveness and treatment response in pediatric brain tumors, supporting their potential for non-invasive disease stratification and monitoring.

## Introduction

Malignant brain tumors are the leading cause of cancer-related death in children and adolescents but early disease detection and close monitoring of treatment efficacy can improve survival [1]. More than 20% of childhood brain tumors are linked to inherited cancer syndromes driven by pathogenic variants in tumor suppressor genes (*DICER1*, *TP53*, *NF1*, *PTCH1*, *SUFU* etc.) [2]. Children with germline *TP53* mutations (Li-Fraumeni syndrome) face a 90% lifetime risk of developing cancer [3] and certain pathogenic variants of *TP53* significantly increase the risk of getting malignant brain tumors during childhood [4]. Early detection of malignant brain tumors can improve survival by enabling complete surgical resection.

New surveillance recommendations suggest that all individuals with brain tumor predisposition syndromes are included in screening programs that utilize magnetic resonance imaging (MRI) of the brain [2]. But MRI is resource-intensive, possesses low sensitivity, and more importantly fails to differentiate true progression from pseudo-progression of the brain tumors [5]. On the other hand, using tissue biopsies to identify disease subtype entails a highly invasive procedure, yet might only capture a static snapshot of the progressive tumor [6]. Furthermore, current targeted therapeutic strategies under development are likely to be more effective, if treatment is initiated early when the disease burden is still low and results in minor symptoms, employing an accurate and reliable method for treatment monitoring. Therefore, there is an urgent unmet clinical need for a more reliable non-invasive diagnosis and prognosis tool that can (i) aid in diagnosis and patient stratification, (ii) identify true disease recurrence, and (iii) indicate response to treatment.

Over the last decade, liquid biopsy approaches, including extracellular vesicles (EVs), have emerged as promising sources of biomarkers. EVs are small membrane-bound vesicles released by cells into the extracellular space and carry proteins, nucleic acids, and lipids reflective of their cell of origin. In cancer, tumor-derived EVs detectable in circulation provide molecular information on disease processes as well as insights into tumor–microenvironment interactions [7, 8]. Because plasma EVs have a short half-life [9–11], they capture the functional disease state in near real time [12]. Collectively, circulating cancer-derived EVs represent a valuable biomarker platform with potential applications in early brain tumor detection, disease monitoring, and assessment of therapeutic response [13].

In a previous study, we showed that EV-associated Syndecan-1 (EV-SDC1) level in plasma is higher in adult patients with glioblastoma compared to patients with low-grade gliomas with an area under the ROC curve of 0.81 [14]. Also, we found strong support of plasma EV-SDC1 originating from glioblastoma tumors, as plasma EV-SDC1 correlated with SDC1 protein expression in matched patient tumors, and plasma EV-SDC1 was decreased postoperatively depending on the extent of surgery [14]. Additionally, publicly available RNA expression data show that SDC1 is upregulated in malignant brain tumors in children [15]. Notably, this gene is particularly upregulated in certain malignant tumor types associated with germline predisposition syndromes (such as sonic hedgehog medulloblastoma [16], embryonal tumor with multilayered rosettes [17]. Given this background and the strong evidence of EV-SDC1 in malignant brain tumors, in this study, we aimed to investigate whether EV-SDC1 can serve as a reliable biomarker for analyzing tumor burden and longitudinal monitoring of disease progression in a well-defined cohort of children and adolescents with brain tumors. EV analysis using advanced microscopy and measurement of SDC1 protein in EVs, we found EV-SDC1 to discriminate high grade (ependymoma, diffuse midline glioma, and atypical teratoid/rhabdoid tumor (AT/RT)) from low grade pilocytic astrocytoma pediatric brain tumor patients.

## Methods

### Study design and patient sample collection

Plasma samples and other material used in this study, were from participants enrolled at the Department of Clinical Medicine – Children and Adolescents, Aarhus University hospital between 2021 and 2025. Inclusion criteria were patients age <18 and newly diagnosed brain tumors. Participants were diagnosed with routine MRI of the brain, surgical and pathological procedures (including immunohistochemistry and methylation analysis (Affymetrix 850K)). Participants with germinoma were diagnosed based on the level of tumor markers such as alpha-fetoprotein and human Chorionic Gonadotropin in the spinal fluid. Patients received standard oncological treatment and were followed according to local and national recommendations as well as repeated MRI examinations when needed. Blood samples were collected in EDTA tubes, centrifuged at 2,000 × *g* for 10 mins at room temperature (RT) and plasma was stored at −80°C. Plasma samples were collected at baseline (preoperative, n=60) and after surgery (postoperative, n=18). The study was carried out according to the ICH/GCP guideline and in agreement with the Helsinki declaration and was approved by the local ethics committee, the National Committee on Health Research Ethics in Denmark (H-20000845).

### EV enrichment

EVs were enriched from plasma samples using sepharose-based SEC columns (35 nm, Gen 2 IZON Science) as previously described [14]. Plasma samples were thawed on ice for the first time after freezing. Five hundred microliter plasma aliquots were applied to the column, and 15 fractions of 500 μL were collected immediately with 2 mmol/L CaCl_2_ in PBS as the elution buffer. Fractions 5 to 10, corresponding to the EV elution profile, were pooled and freeze-dried using Mini Lyotrap (LTE Scientific). Aliquots of pooled intact EVs were used for EV characterization. For SDC1 quantification, the EV pellet was lysed with cell lysis buffer II (FNN0021, Invitrogen) as per manufacturer’s instructions. Lysed EV proteins were desalted using PD-10 columns (Cytiva) according to the standard protocol and purified EV proteins were concentrated by freeze-drying and stored at −80°C until used in SDC1 ELISA.

### EV characterization

#### Cryo-Transmission Electron Microscopy (cryo-TEM)

The intact EV samples used for cryo-TEM underwent no more than one freeze-thaw cycle and were thawed immediately prior to grid preparation. A 3 µL aliquot of the EV suspension was applied to glow-discharged, lacey carbon-coated 200 mesh copper grid (Quantifoil). Using a back-blotting approach, 1 µL of ultrapure water was added to the reverse (non-carbon) side before blotting for 3 s with filter paper. The grids were then rapidly vitrified by plunging into liquid ethane cooled to approximately −184°C. Vitrified grids were stored in liquid nitrogen until imaging. Data were collected on a Titan Krios G3i transmission electron microscope (Thermo Fisher Scientific) equipped with a K3 direct electron detector and a BioQuantum energy filter (Gatan). Images were recorded at a nominal magnification of 42,000×, corresponding to a physical pixel size of 2.1 Å/pixel, with an acceleration voltage was 300 kV. ImageJ software (1.53t) was used to add scale bars to the acquired micrographs.

#### Immunoblotting

Dot blots were performed by spotting 3 µL of sample on nitrocellulose membranes (Thermo Fisher) with a 0.45 μm pore size. Membranes were dried for 30 mins before blocking in 5% BSA in PBS for one hour.

The blots were then incubated with either a monoclonal antibody targeting CD63 (10628D, Invitrogen), a monoclonal antibody targeting CD9 (MA1-19301, Invitrogen) or a monoclonal antibody targeting SDC1 (MI15, Stemcell technologies) overnight at 4°C. The blots were then washed 3x PBS+0.05% Tween-20 followed by incubation with HRP-conjugated anti-mouse secondary antibody (31432, Thermo Fisher). Blots were washed 3× in PBS+0.05% Tween-20 prior to development using Clarity Western ECL (BioRad) and visualization using a Chemidoc Imager (BioRad).

Using ImageJ (1.51t), the intensity of the dot blot was quantified by determining the integrated density of each dot after background correction with a rolling ball radius set to 25 pixels.

#### NanoFlow Cytometric analysis

The size distribution and particle concentration of EVs were measured by NanoFlow cytometry (CytoFLEX Nano, Beckman Coulter, Souzhou, China). The instrument was calibrated for particle measurements using CytoFLEX nano QC Scatterspheres (Beckman Coulter, C93495-AA) and CytoFLEX nano QC Fluorospheres (Beckman Coulter, C93496-AA) as well as Nanoscale Sizing Standards (Beckman Coulter, D03231). From representative samples, 20,000 events were collected [18]. The size distribution and particle concentration were analyzed using CytExpert nano software (v. 1.1.0.6, Beckman Coulter, Souzhou, China).

### EV-SDC1 Quantification

#### NanoFlow cytometry

EVs from representative samples were identified as expressing CD63 (MA5-18149, Invitrogen, A488) or SDC1 (17-1389-42, Invitrogen, APC) on the CytoFlex. From representative sample, 1 µL of EVs were incubated with 1 µL of each antibody and 3 µL PBS, resulting in a total of 2 µL of antibodies and a dilution of 1:5 of the EVs. The mixture was then incubated for 30 mins at 37°C. Next, 1 μL of the EV/antibody mix was diluted 1:1000 in PBS, resulting in the EV samples being diluted 1:5000.

Moreover, one sample was incubated for 30 mins at 37°C with the following: 4 μL of PBS or 1 μL of a single undiluted antibody plus 3 μL of PBS, serving as controls. PBS alone and PBS plus a mix of antibodies were also included as controls. The samples and controls were run on the CytoFlex at a controlled flow of 1 μL/min.

The data was analyzed using FlowJo^TM^ software v. 10.8 (BD Life Sciences) with the following stepwise gating strategy applied to the samples. Based on a PBS-only control, background events from the buffer solution were gated out (VSSC1-H). Next, doublets were excluded using the VSSC1-width vs VSSC1-H. To ensure that only EVs positive for either CD63 or SDC1, a conservative gating strategy was applied, where gates were defined solely based on the unstained sample and the buffer control (Fig S1) to exclude noise and instrument background. The antibody mix control was used exclusively to correct for signal originating from free antibodies in solution. Specifically, the signal measured in the antibody mix was subtracted from the corresponding stained samples, thereby reducing the contribution of unbound antibody and isolating the true EV-associated CD63 and SDC1 signals.

#### Total internal reflection fluorescence (TIRF) microscopy

TIRF microscopy was performed using an Oxford Nanoimager (ONI) microscope. EV samples were incubated with the SDC1 antibody (17-1389-42, Invitrogen, APC) for 30 mins at 37°C. Subsequently, the EV/antibody mix were membrane-labeled and captured on a passivated surface using an established method [19]. In brief, EVs were mixed with 300 DOPE-ATTO 488 (ATTO-TEC) and 150 biotinylated lipid-anchors (DSPE-PEG(2000)biotin, ATTO-TEC)) per EV in PBS and incubated for 30 mins at 37°C under gentle shaking at 300 rpm. The biotinylated and fluorescently labeled EVs were then captured on a passivated glass slides with a microscope chamber (sticky-Slide VI 0.4, ibidi). Biotinylated, fluorescently labeled EVs were immobilized on passivated glass slides mounted with microscope chambers as described previously [20] (Sticky-Slide VI 0.4, Ibidi). Glass slides were cleaned by sequential sonication (10 min each) in 2% Helmanex, Milli-Q water, and methanol, followed by plasma activation. The activated surfaces were assembled with Sticky-Slide chambers and functionalized with BSA–biotin for 30 mins. After washing, neutravidin (0.1 g/L) was added to enable specific EV capture. Excess reagents were removed by thorough PBS washing. The EV/antibody mix were added to the surface and allowed to bind for 5 mins, before being washed away using 2 mL PBS. The data was acquired with a 100× 1.4 NA oil immersion objective and an exposure time of 100 ms. Approximately 162 images were acquired for each condition. Images in two channels were taken with alternating laser excitation for each field of view, with the 488 nm laser at 7% and the 640 nm laser at 15%. One field of view corresponds to a physical field of view of 50 μm x 80 μm. For control sample EV without antibody were imaged.

TIRF microscopy data were analyzed using an in-house developed automated python-based image analysis. Spots corresponding to EVs were detected as described before. The membrane channel was used for spot detection and the background-corrected signal for the respective spots was extracted from both channels (membrane and antibody). For the negative control, EVs without antibody staining were detected and the corresponding antibody signal was extracted from the control sample containing antibody in PBS without EVs.

#### ELISA

SDC1 levels in plasma EV samples were analyzed using the Human SDC1 ELISA Kit (EHSDC1, Thermo Scientific) according to the manufacturer’s instructions. Briefly, 100 μL of each standard and lysed EV samples were added to appropriate wells and incubated for 2.5 hour at RT with gentle shaking. After washing, 100 μL of 1× biotinylated antibody was added and incubated for 1 hour at RT. Following another wash, 100 μL of streptavidin–HRP solution was added to each well and incubated for 45 mins at RT. One hundred microliters of TMB substrate was then added to samples for 30 mins at RT in the dark with gentle shaking, followed by the addition of 50 μL of stop solution. The absorbance was measured at 450 nm and 550 nm in a spectrophotometer (FLUOstar OPTIMA).

### Tumor volume analysis

Tumor volume measurements from baseline and postoperative patients were directly obtained from MRI scans acquired as part of the patients’ routine diagnostic and follow-up procedures.

Tumor volume (*V*) was estimated using the formula:

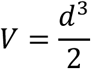

where *d* represents the maximum tumor diameter measured on MRI.

### Bioinformatics and statistical analyses

The pediatric brain tumor subtype data sets for SDC1 mRNA expression were obtained from the Open Pediatric Brain Tumor Atlas (OpenPBTA) portal [21], established in 2018 by the Children’s Brain Tumor Network (CBTN, https://cbtn.org) and the Pacific Pediatric Neuro-Oncology Consortium (PNOC, https://pnoc.us). *P*<0.05 was considered statistically significant. Data are presented as the means ± SD. All figures were prepared and analyzed using GraphPad Prism (version 10.0).

### Results

### Patient description

In total, 102 participants were screened and found eligible to participate in the study, from which a baseline cohort (n=60) and a longitudinal cohort (n=18) were recruited. An outline of the patient characteristics including different tumor subtypes and grades, demographics, and mutations is given in Table S1 and a flow chart outlining inclusion, and exclusion criteria is shown in Fig 1.

**Figure 1.**
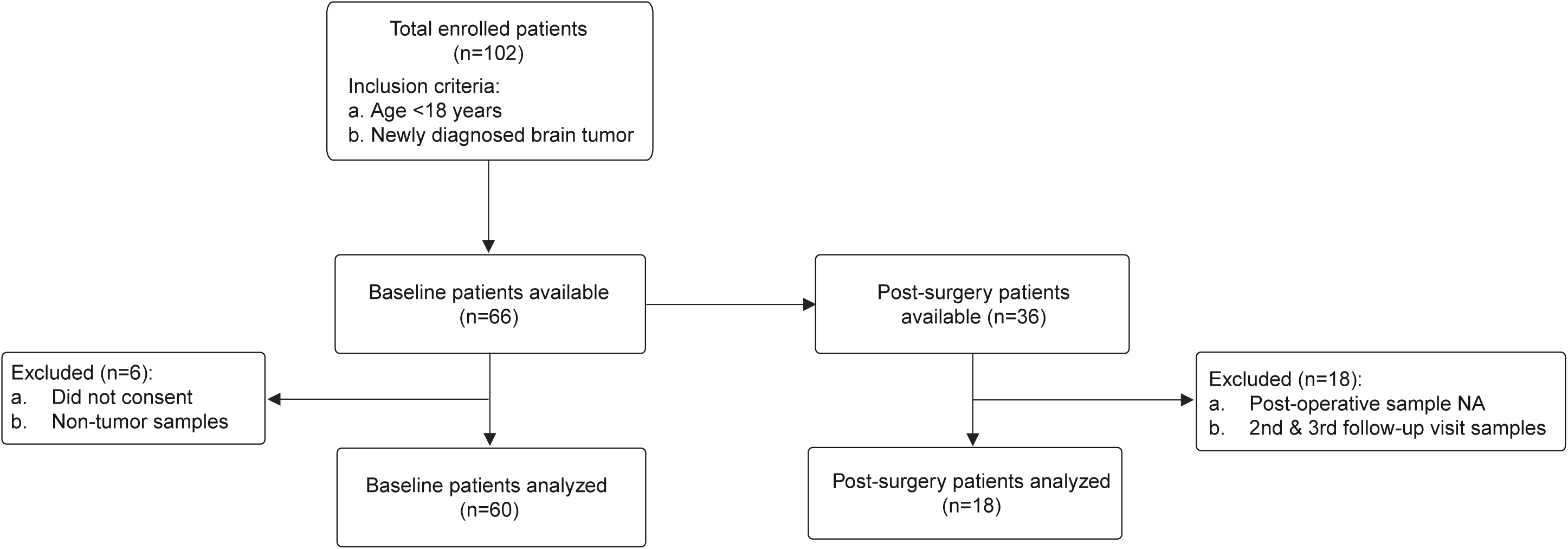
Consort diagram showing pediatric brain tumor patient cohort, inclusion and exclusion criteria. NA, not available.

### Characterization of EVs and SDC1^+^ EVs from pediatric brain tumor patients

EVs isolated from pediatric brain tumor patients by SEC were extensively characterized by immunoblotting, NanoFCM, Cryo-TEM, and TIRF (Fig 2A). The size distribution of plasma-derived EVs was assessed by NanoFlow cytometry which displayed the expected features characteristic of small EVs. The median particle size was 78 nm and the average particle concentration in representative samples was 4.2 x 10^11^ particles/mL (CI: 3.3 x 10^11^ – 5.1 x 10^11^ particles/mL) (Fig 2B). We further corroborated the nature of EVs by Cryo-TEM that revealed a heterogenous population of spherical, lipid-bilayer enclosed vesicles with diameters primarily ranging from 50-150 nm. We observed intact membrane delimited vesicles with minimal aggregation or structural collapse, confirming preservation of vesicular integrity following SEC-based isolation and lyophilization (Fig 2C). The presence of EV-associated canonical markers, CD9 and CD63, was established by dot blot immunoassays, verifying enrichment of vesicular material (Fig 2D & Fig S2). Notably, plasma-derived EVs from patients with different tumor grades had detectable levels of SDC-1, as analyzed by dot blot immunoassays (Fig 2D & Fig S2) and independently verified by NanoFlow cytometry (Fig S3). A maximum of 1% of the detected events were positive for SDC1, as determined with NanoFlow cytometry (Fig 2E & Fig S3). More importantly, single-EV imaging by TIRF microscopy confirmed the recruitment of SDC1 at the individual vesicle level. Fluorescently labeled SDC1 antibodies colocalized with membrane-stained EVs, validating the NanoFlow cytometry findings and supporting that SDC1 is displayed on the surface of a defined EV subset (Fig 2F & G). The proportion of SDC1-positive EVs observed by TIRF microscopy corresponded closely with the NanoFlow cytometry measurements, highlighting methodological consistency across single-particle detection platforms.

**Figure 2.**
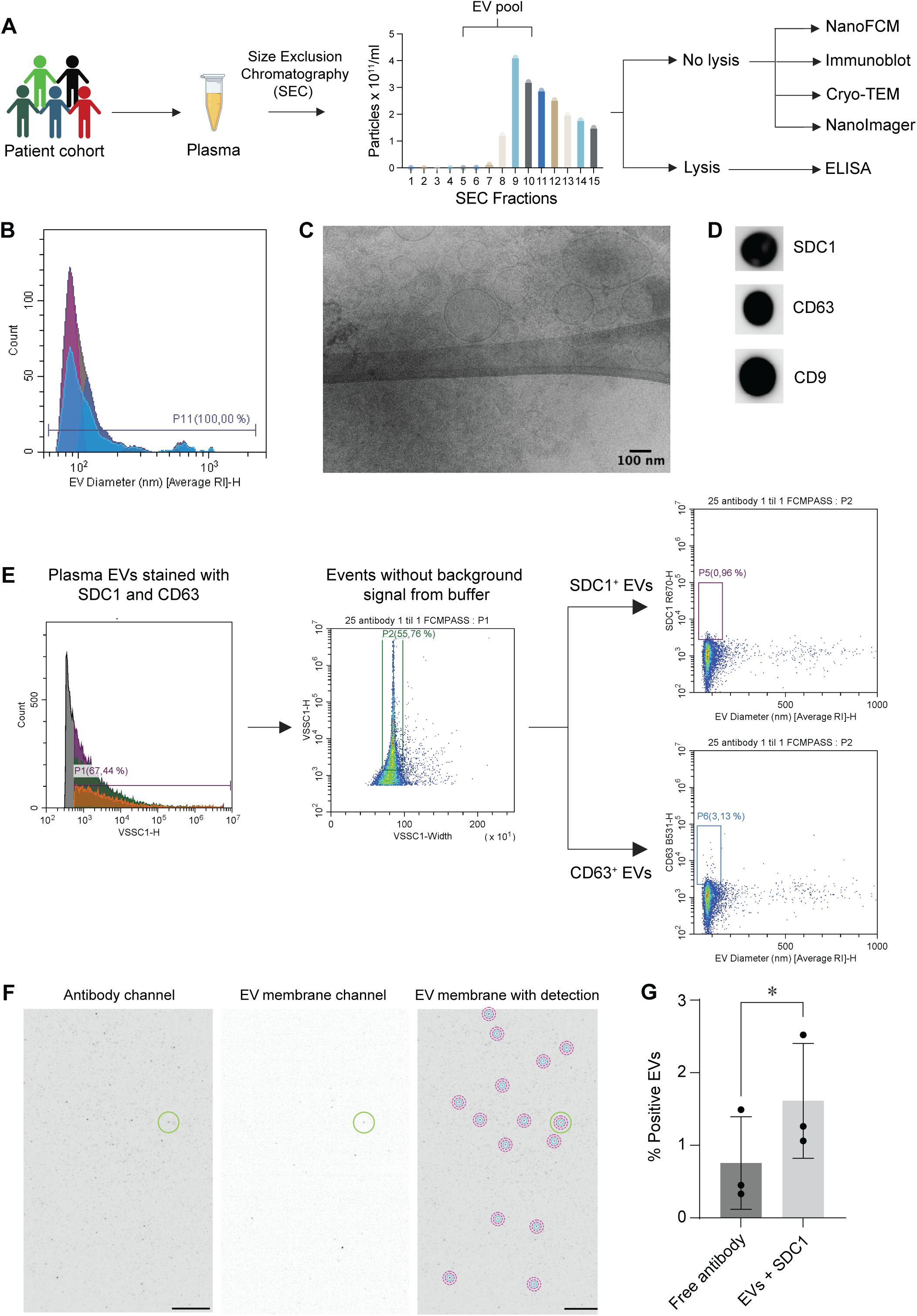
Molecular characterization of extracellular vesicles (EVs). (A) Schematic overview of experimental workflow, including EV isolation and downstream characterization. (B) Particle size distribution of EVs measured by NanoFlow cytometry, showing a dominant population of particles within the expected size range. (C) Electron microscopy image of EVs displaying characteristic vesicular morphology and size. Scale bar as indicated. (D) Immunoblot analysis confirming enrichment of canonical EV associated markers. (E) NanoFlow gating strategy illustrating exclusion of background events, removal of doublets and finally identification of SDC1^+^ and CD63^+^ EVs populations, respectively. (F) Total Internal Reflection Fluorescence (TIRF) representative image showing from left to right; Antibody signal, EV signal and merged channels highlighting EV membrane associated region of interest detection for signal integration and local background correction. Scale bar = 10 µm (G) Quantification of TIRF microscopy data confirming the presence of SDC1^+^ EVs.

Collectively, these findings confirmed that SEC-based enrichment yielded a pure population of structurally intact small EVs suitable for downstream quantitative analyses of SDC1 expression.

### EV-SDC1 is associated with pediatric brain tumor grades

In a previous study, we demonstrated that EV-associated SDC1 levels correlate with tumor grade in adult glioma patients [14]. Building on these findings, we measured EV-SDC1 levels by ELISA in a well-defined and heterogeneous cohort of baseline pediatric brain tumor patients (n=60) (Fig 3A & Table S2), aiming to investigate the potential association between EV-SDC1 and tumor grade in this population. For comparison, all patient tumor grades were categorized from 1-4 as per WHO 2021 CNS tumor guidelines [22]. Consistent with the tumor heterogeneity, substantial variability in EV-SDC1 levels was observed both within and across different tumor grades (Fig 3A&B). Grade 3 tumors (n=6, including germinoma, high grade glioma/astrocytoma, and ependymoma) had significantly higher EV-SDC1 levels compared to patients with grade 1 pilocytic astrocytoma (p=0.02) and grade 2 tumors (p=0.06) (Fig 3B). Next, we evaluated the diagnostic accuracy of EV-SDC1 as a potential marker to discriminate grade 3 from grade 1 and grade 2 patients, which showed an area under the receiver operating characteristic (AUROC) curve; grade 3 vs grade 1, AUROC of 1.00 (CI: 1.00–1.00, p=0.018) and grade 3 vs grade 2, AUROC of 1.00 (CI: 1.00–1.00, p=0.039) (Fig 3C, E & G), indicating the potential of EV-associated SDC1 to discriminate grade 3 from low grade 1 and grade 2 patients. We also observed a similar trend of high EV-SDC1 levels in grade 4 glioblastoma compared to grade 1 and grade 2 patients, albeit not significant; grade 1 vs grade 4 (p=0.27; AUROC=0.62, p=0.2) and grade 2 vs grade 4 (p=0.74; AUROC=0.55, p=0.76) (Fig 3B, D, F & G).

**Figure 3.**
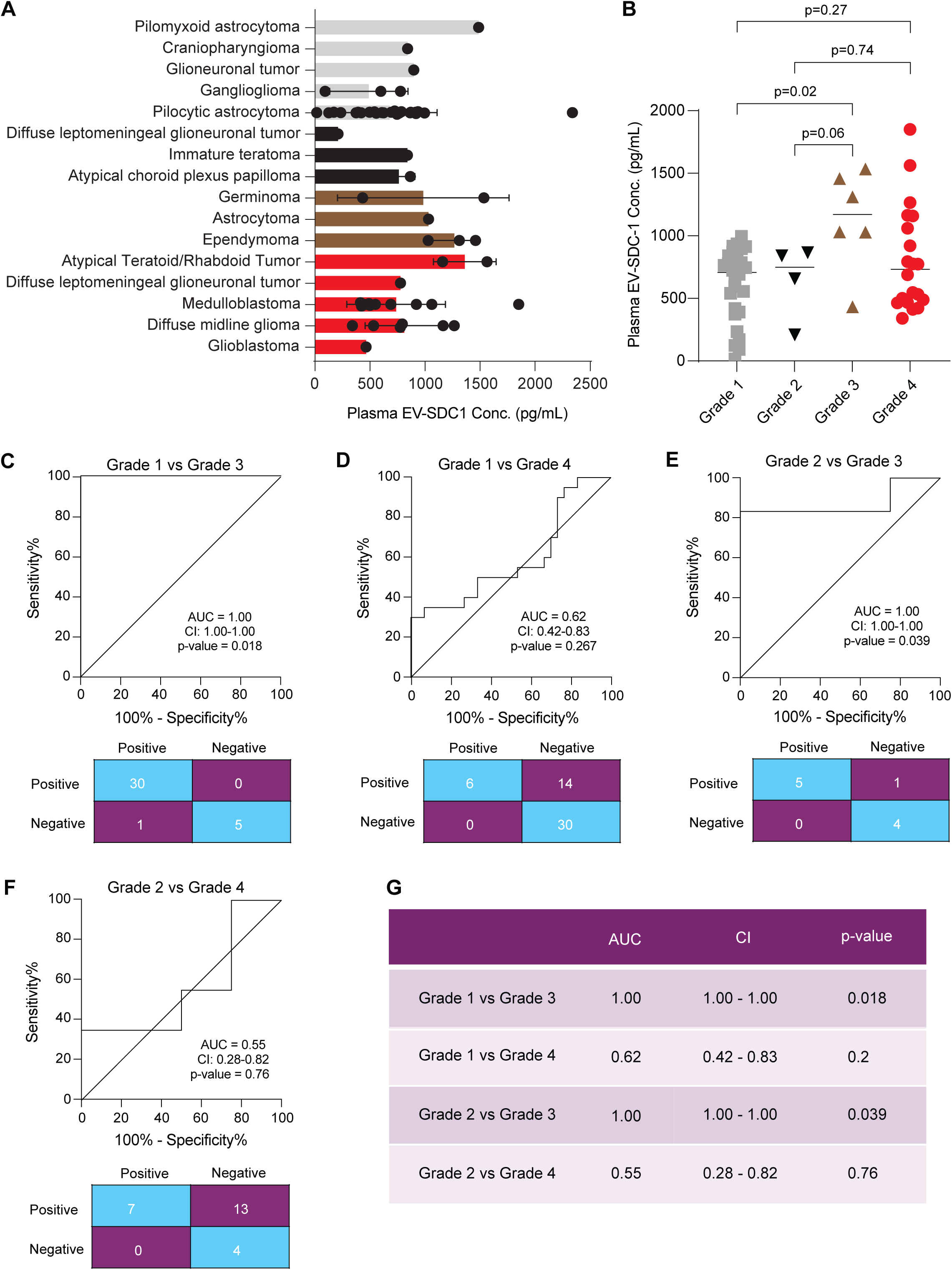
Differential levels and diagnostic accuracy of extracellular vesicle syndecan 1 (EV-SDC1) in pediatric brain tumor subtypes. (A) Bar graph showing EV-SDC1 levels across different pediatric brain tumor subtypes. (B) Scatter plot displaying differential levels of EV-SDC1 as per WHO 2021 recommended grades 1-4. Receiver Operating Characteristic (ROC) curve and confusion matrix revealing diagnostic accuracy of EV-SDC1 in distinguishing (C) Grade 1 vs Grade 3 patients, (D) Grade 1 vs Grade 4 patients, (E) Grade 2 vs Grade 3 patients, and (F) Grade 2 vs Grade 4 patients. (G) Table showing compilation of ROC area under the curve, confidence interval (CI), and p-values. P-values <0.05, significant.

Together, these findings indicate the potential of EV-SDC1 as an individual biomarker to discriminate high grade from low grade, particularly grade 1 pilocytic astrocytoma, brain tumor patients.

### Tumor SDC1 mRNA levels can distinguish pediatric brain tumor patients

Following observations that EV-SDC1 levels were elevated in high-grade compared to low-grade pediatric brain tumor patients, we sought to validate these findings at the transcriptional level. We investigated the SDC1 mRNA expression by accessing the large cohort of pediatric brain tumor patients (n=337) including low-grade pilocytic astrocytoma (n=105) and high-grade subtypes including ependymoma (n=59), medulloblastoma (n=91), high grade glioma (HGG)/astrocytoma (n=32), AT/RT (n=23), and diffuse midline glioma (n=27) from OpenPBTA (Fig 4A & Table S3). The SDC1 mRNA expression across different grades revealed a similar pattern of variation to EV-SDC1 consistent with the heterogeneous nature of pediatric brain tumors (Fig 4A). Expectedly, SDC1 levels were higher in high-grade tumors including ependymoma, HGG/astrocytoma, medulloblastoma, AT/RT, and diffuse midline glioma compared to low-grade pilocytic astrocytoma patients; ependymoma vs pilocytic astrocytoma (p<0.001); HGG/astrocytoma vs pilocytic astrocytoma (p<0.001); medulloblastoma vs pilocytic astrocytoma (p=0.365); AT/RT vs pilocytic astrocytoma (p<0.001); and diffuse midline glioma vs pilocytic astrocytoma (p<0.001) (Fig 4A). When individual high grade tumor subtypes were compared to grade 1 tumors, we observed SDC mRNA could discriminate ependymoma vs grade 1 with a diagnostic accuracy (AUROC=0.71; p<0.001) (Fig 4B & G), HGG/astrocytoma vs grade 1 (AUROC=0.71; p<0.001) (Fig 4C & G), medulloblastoma vs grade 1 (AUROC=0.61; p=0.006) (Fig 4D & G), AT/RT vs grade 1 (AUROC=0.95; p<0.001) (Fig 4E & G), and diffuse midline glioma vs grade 1 (AUROC=0.84; p<0.001) (Fig 4F & G).

**Figure 4.**
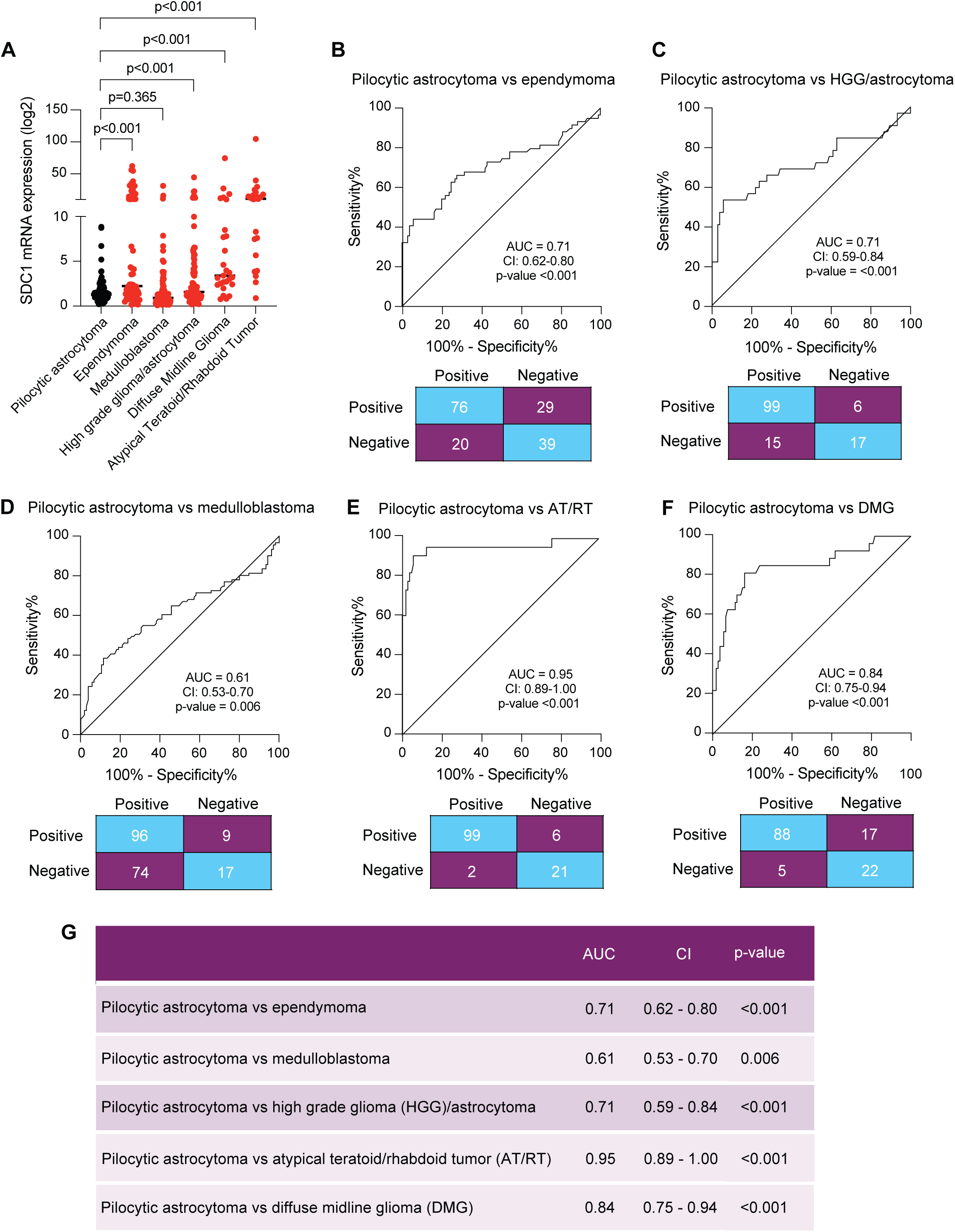
Differential levels and diagnostic accuracy of syndecan 1 (SDC1) mRNA in pediatric brain tumor subtypes, data retrieved from Open Pediatric Brain Tumor Atlas (OpenPBTA) platform. (A) Bar graph displaying SDC1 mRNA levels across different pediatric brain tumor subtypes including pilocytic astrocytoma, ependymoma, medulloblastoma, high-grade glioma (HGG)/astrocytoma, diffuse midline glioma, and Atypical Teratoid/Rhabdoid Tumor (AT/RT). Receiver Operating Characteristic (ROC) curve and confusion matrix showing diagnostic accuracy of SDC1 mRNA in distinguishing (B) Pilocytic astrocytoma vs ependymoma, (C) Pilocytic astrocytoma vs HGG/astrocytoma, (D) Pilocytic astrocytoma vs medulloblastoma, (E) Pilocytic astrocytoma vs AT/RT, and (F) Pilocytic astrocytoma vs diffuse midline glioma pediatric brain tumor patients. (G) Table showing compilation of ROC area under the curve, confidence interval (CI), and p-values. P-values <0.05, significant.

Collectively, these findings further confirm the potential of SDC1 as a marker to identify high grade vs low grade pediatric brain tumor patients and indirectly support the role of brain tumor tissues as the source contributing to the increased level of circulating EV-SDC1.

### EV-SDC1 can predict response of pediatric brain tumor patients to treatment

To understand if EV-SDC1 levels can predict the tumor burden in postoperative patients, we examined the change in the levels of EV-SDC1 in pediatric brain tumor patients (n=18) before and after surgery by ELISA (Fig 5A & B, Table S4). Patients included both low-grade pilocytic astrocytoma and high-grade tumors such as ependymoma, diffuse midline glioma, and medulloblastoma. Overall, 8 out of 18 baseline patients (∼45%) showed a reduction in EV-SDC1 levels post-surgery (Fig 5 A&B, indicated in green) and increased levels of EV-SDC1 were observed in the remaining patients (n=10, indicated in red). Upon closer look, it was revealed that most of the patients with elevated EV-SDC1 levels post-surgery had only a partial resection of their tumors (Fig 5B). The presence of residual tumor and the longer time to follow-up visit after surgery indicate tumor relapse that is reflected in the increased EV-SDC1 levels.

**Figure 5.**
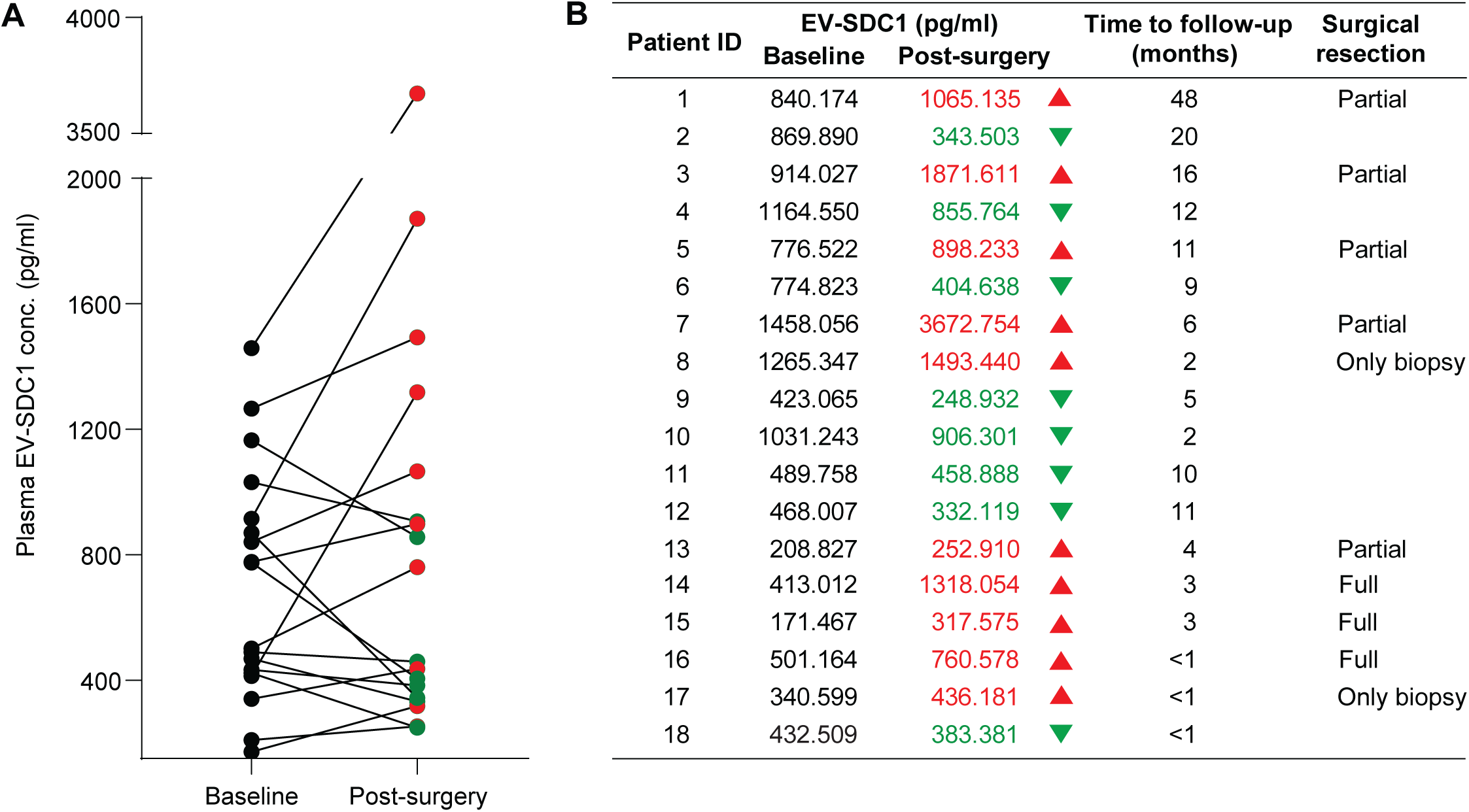
Analysis of extracellular vesicle-syndecan 1 (EV-SDC1) as marker of tumor burden in pediatric brain tumor subtypes. (A) Spaghetti plot displaying the relative changes in the level of EV-SDC1 in baseline (before surgery) and after surgery in pediatric brain tumor patients. (B) Table showing EV-SDC1 values before and after surgery, time to first follow-up, and surgical status (partial or full resection) of pediatric brain tumor patients.

Given the compelling diagnostic accuracy of EV-SDC1 to identify high grade pediatric brain tumors from low grade pilocytic astrocytoma patients and its ability to predict the tumor burden after surgery, we sought to examine the correlation of EV-SDC1 with tumor volume. Measurements of tumor volume in baseline as well as postoperative patients (n=40) were performed using MRI images (Fig S4A). We did not observe a significant correlation between EV-SDC1 levels and tumor volume in baseline and post-surgery patients (*r*=0.14, p=0.4) (Fig S4B) indicating that EV-SDC1 levels vary depending on tumor severity rather than changes in the volume of the tumor.

## Discussion

In this study, we identified and characterized a small subpopulation of circulating EVs carrying SDC1 and demonstrated that the abundance of these vesicles correlates with tumor grade in pediatric glioma patients. Using complementary single-EV and bulk approaches, we confirmed by both nano flow cytometry and TIRF microscopy, that SDC1 is expressed on only a minor fraction of plasma EVs, corresponding to approximately 1% of the total EV plasma population. Quantification by ELISA further showed that plasma EV-SDC1 levels were significantly higher in patients with high-grade tumors compared to those with low-grade disease, suggesting that EV-SDC1 reflects tumor aggressiveness. Together, these findings provide evidence that EV-SDC1 may serve as a minimally invasive biomarker with potential value for monitoring disease progression in patients with pediatric brain tumors.

SDC1 is a transmembrane heparan sulfate proteoglycan that can regulate cell adhesion, migration and interactions with the extracellular matrix [23]. Aberrant overexpression of SDC1 has been reported in numerous malignancies, including multiple myeloma, breast cancer, pancreatic ductal adenocarcinoma, colorectal cancer, prostate cancer, and hepatocellular carcinoma, where it promotes tumor cell invasion, angiogenesis, immune evasion, and resistance to chemotherapy and targeted therapies [24]. SDC1 can be released into the extracellular environment either as a soluble ectodomain following proteolytic cleavage [25] or via packaging into extracellular vesicles [14]. The detection of SDC1 on plasma EVs in our study indicates that, SDC1 is sorted into vesicular membrane structures.

EVs were enriched using size-exclusion chromatography, reducing contamination from soluble plasma proteins. Cryo-EM and nano flow cytometry confirmed that the enriched vesicles displayed the expected morphology and size distribution of small EVs, while immunoblotting verified the presence of canonical tetraspanins CD9 and CD63. Importantly, both nano flow cytometry and TIRF microscopy revealed similar frequencies of SDC1-positive vesicles, highlighting the reproducibility of SDC1 detection across different analytical modalities.

The biological implications of SDC1-positive EVs merit further exploration. Given SDC1’s capacity to bind growth factors, cytokines, and extracellular matrix components through its heparan sulfate chains, EVs carrying SDC1 could act as mobile signaling platforms within the tumor microenvironment. They may facilitate angiogenesis, modulate immune responses, or promote pre-metastatic niche formation by transferring heparan-bound ligands to recipient cells.

Given the ability of EV-SDC1 to reflect pediatric brain tumor aggressiveness, we expected the levels of EV-SDC1 to be increased in ependymoma, medulloblastoma, and glioblastoma that are classified as high-grade tumors according to WHO 2021 CNS Tumor classification system. However, we observed a relatively lower level of EV-SDC1 in patients with medulloblastoma, which could have resulted from the non-glial embryonal origin of the tumor unlike the glial origin of ependymoma, diffuse midline glioma, and high-grade glioblastoma.

In this patient cohort, we did not observe a consistent and significant correlation between high EV-SDC1 levels in ependymoma, medulloblastoma, and glioblastoma and their volume. Similarly, there was no correlation between residual tumor volume in postoperative patients and their EV-SDC1 levels. This could have resulted partly from the fact that tumors are highly heterogeneous in nature and SDC1 is only expressed on cells of glial origin and not embryonal.

Several technical and biological considerations should be acknowledged. The overall abundance of SDC1-positive EVs was low, which imposes challenges for detection sensitivity and quantification. Although the SEC-based isolation approach effectively reduces plasma protein contamination, residual soluble SDC1 cannot be completely excluded as a contributor to ELISA signal. Moreover, the number of paired pre- and post-operative samples was limited, and larger longitudinal cohorts are needed to validate the observed trends.

Despite these limitations, this work provides novel evidence that SDC1 is selectively displayed on a small subset of circulating EVs in pediatric glioma patients and that its abundance correlates with tumor grade. The consistent and significant detection of SDC1 across single-EV and bulk assays underscores the reliability of the finding and establishes a methodological framework for future biomarker studies. The observation that EV-SDC1 levels fluctuate with surgical intervention and disease progression raises the possibility that this marker could serve as an early indicator of therapeutic efficacy or recurrence. Given the promising observation that EV-SDC1 convincingly identifies high grade ependymoma, diffuse midline glioma, and AT/RT from pilocytic astrocytoma, future work should focus on validating EV-SDC1 as a prognostic and monitoring biomarker in larger, longitudinal cohorts and on elucidating the molecular mechanisms governing SDC1 sorting into EVs.

In conclusion, our findings demonstrate that circulating SDC1-positive EVs represent a distinct vesicle population associated with glioma malignancy. By linking SDC1 levels in vesicles with clinical outcomes, this study advances our understanding of how tumor-derived EVs reflect underlying pathological processes and supports the potential of EV-SDC1 as a minimally invasive biomarker for pediatric glioma diagnosis and disease monitoring.

## Data Availability

All data produced in the present study are available upon reasonable request to the authors

## Acknowledgements

This work was supported by BETA.HEALTH grant (BETA.05 2023-1534, An innovation platform sponsored by Novo Nordisk Foundation) to V.I.C. T.S.M is grateful to Børne Hjernecancer Fonden grant 2021 (Child Brain Tumor Foundation) for supporting this study. S.G. acknowledges Indian Council for Medical Research (ICMR) for ICMR-International Fellowship (INDO/FRC/452/S-76/2023-24-IHD). P.N. is thankful to support from Independent Research Fund Denmark (3105-00153B). The CytoFLEXnano was a generous gift from Carlsberg Foundation (grant CF23-1020). We acknowledge support from the FACS Core Facility at Aarhus University.

## Author contributions

Conceived, T.S.M., and V.I.C.; Conceptualization, V.I.C.; Methodology, J.K.H., M.G.M., and V.I.C.; Investigation and formal Analyses, J.K.H., J.E.J., B.W., M.Z., H.W., S.G., and V.I.C.; Resources, P.N., T.S.M., and V.I.C.; Writing – Original Draft, J.K.H., and V.I.C.; Writing – Review & Editing, J.K.H., J.E.J., B.W., M.Z., H.W., M.G.M., K.A.H., S.G., P.N., T.S.M., and V.I.C.; Supervision, T.S.M., and V.I.C.; Project Administration, V.I.C.; Funding Acquisition, T.S.M., and V.I.C.

---

**Figure S1.**
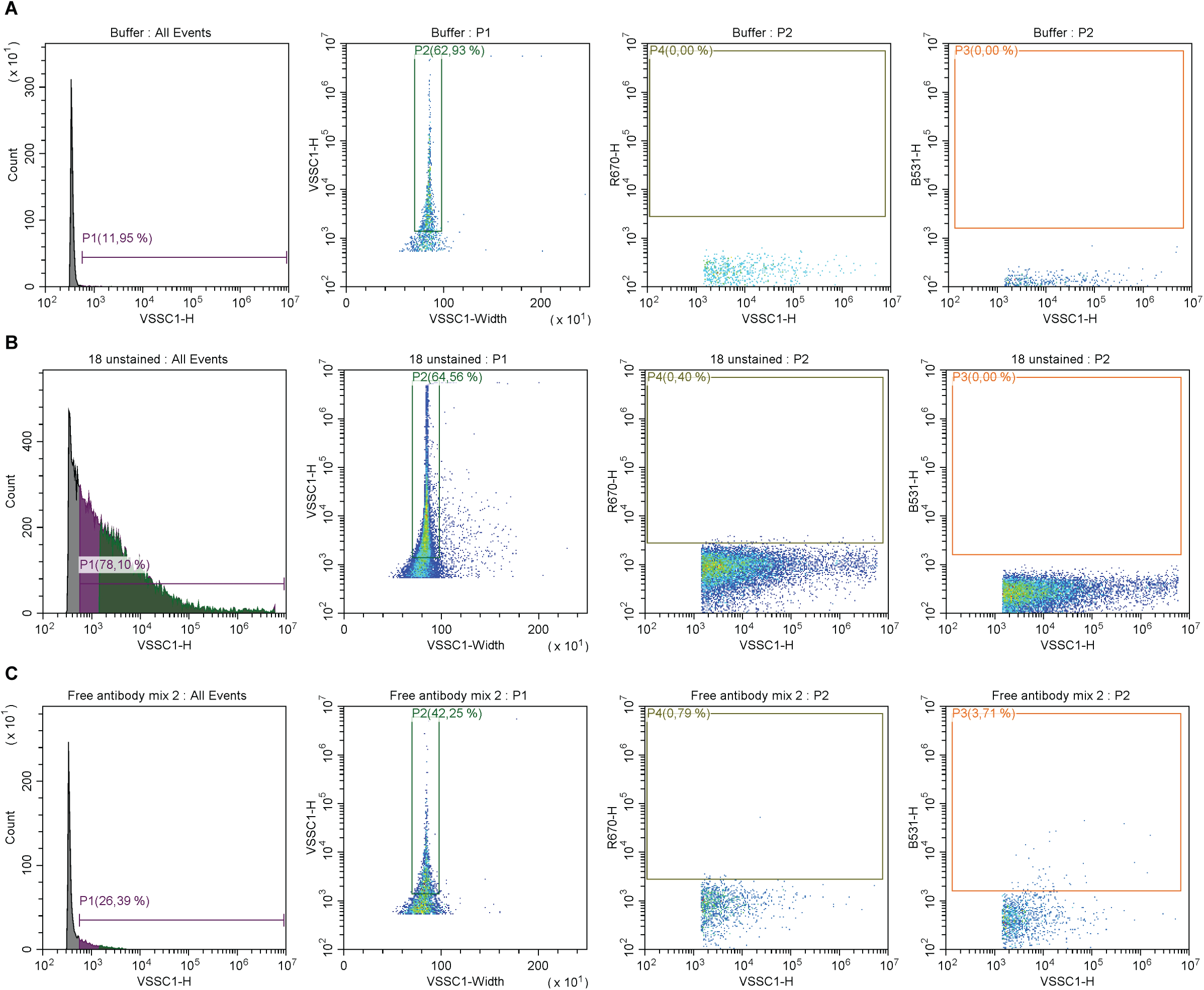
Syndecan 1 (SDC1) and CD63 gating strategy. (A) Buffer (PBS) used for gating background. (B) Unstained sample used for gating.(C) Free mix on antibodies used for estimating free antibodies.

**Figure S2.**
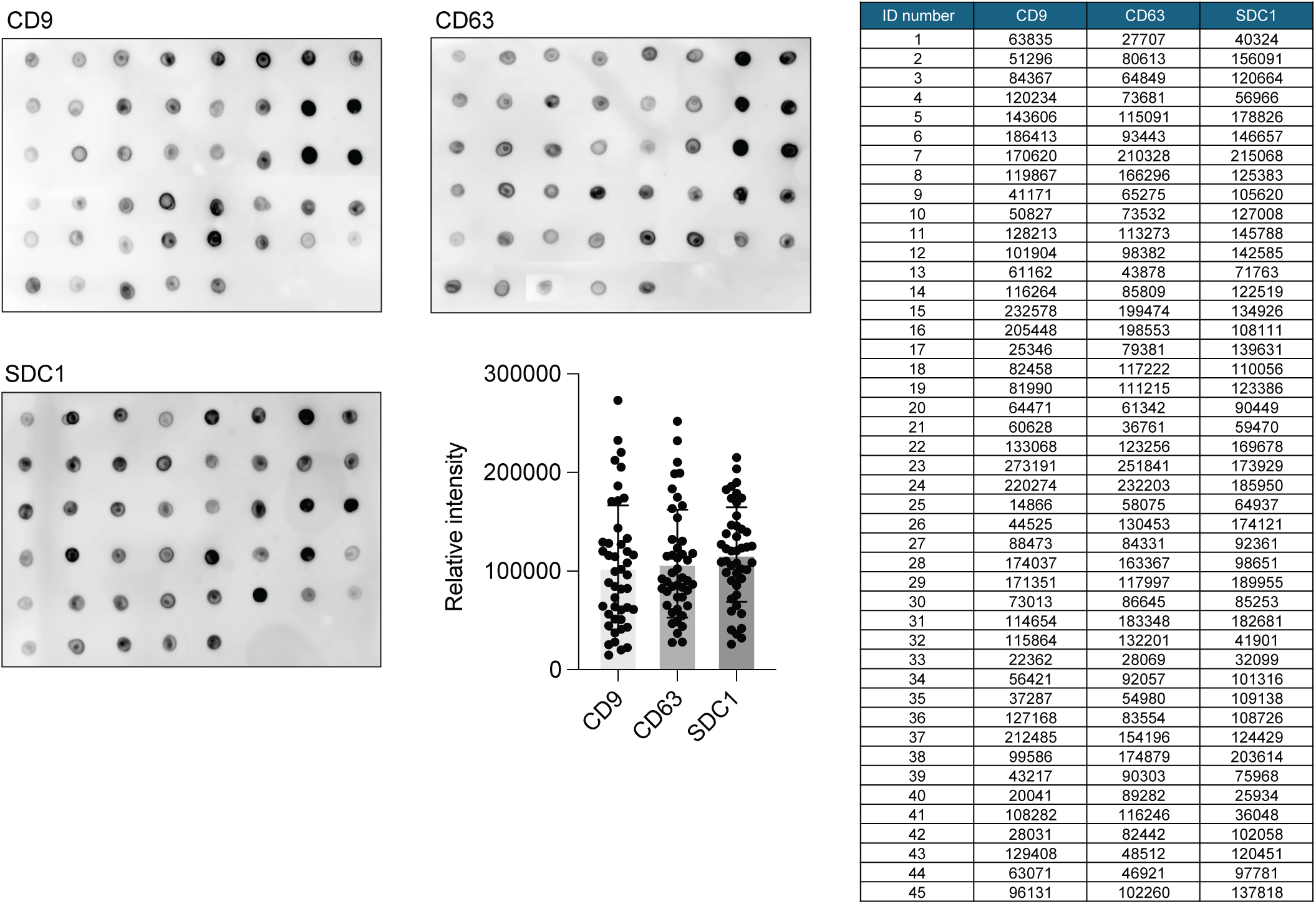
Heterogeneous CD9, CD63 expression and CD63/ syndecan 1 (SDC1) ratios across individual extracellular vesicle (EV) samples. (A) Dot blot analysis of CD9, CD63 and SDC1 expression in EV preparations from 45 individual samples, with representative arrays shown for each marker and corresponding quantification of spot intensities.

**Figure S3.**
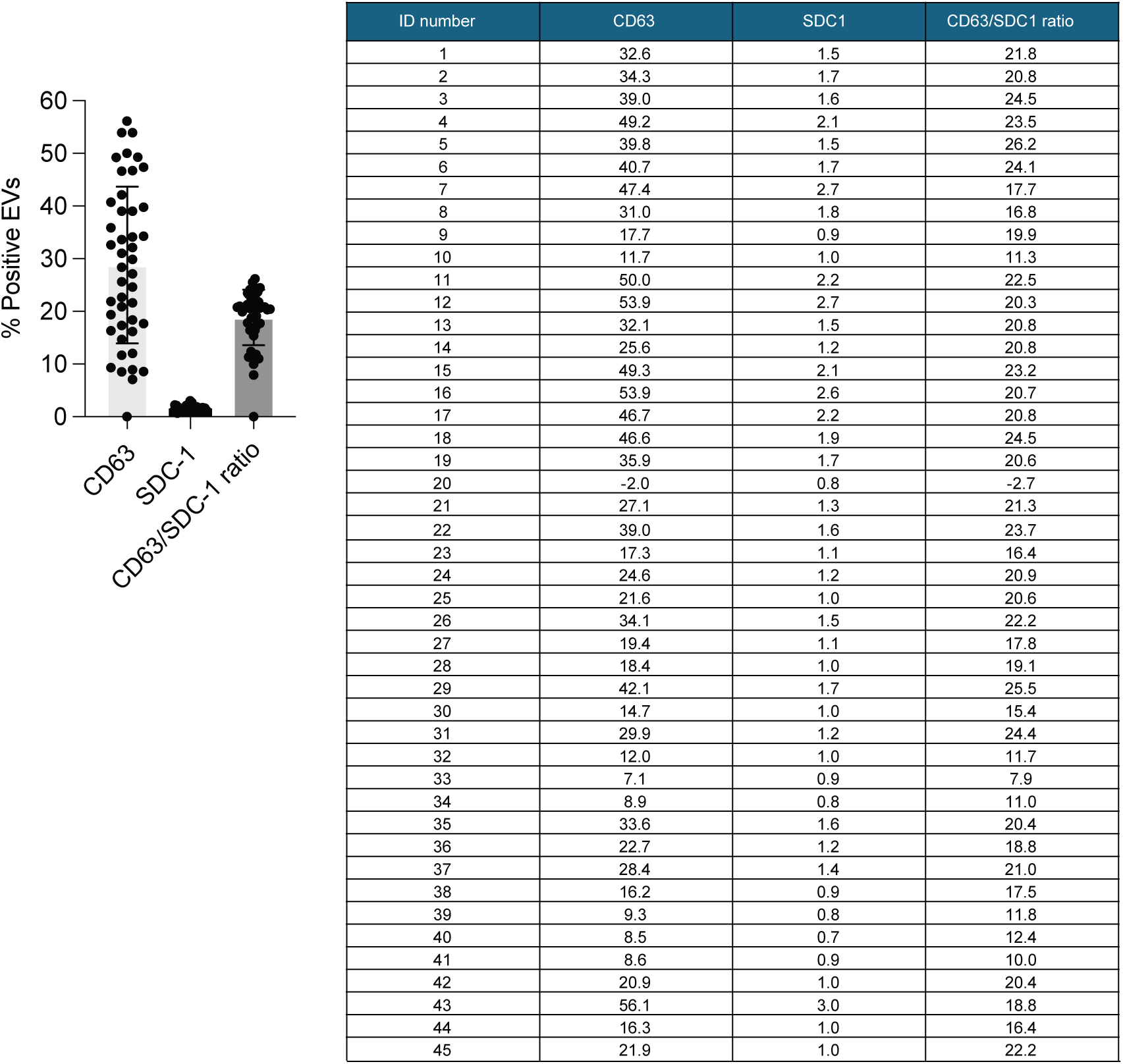
NanoFlow cytometry showing the percentage of CD63- and syndecan 1 (SDC1)-positive extracellular vesicles (EVs) and calculated CD63/SDC1 ratios.

**Figure S4.**
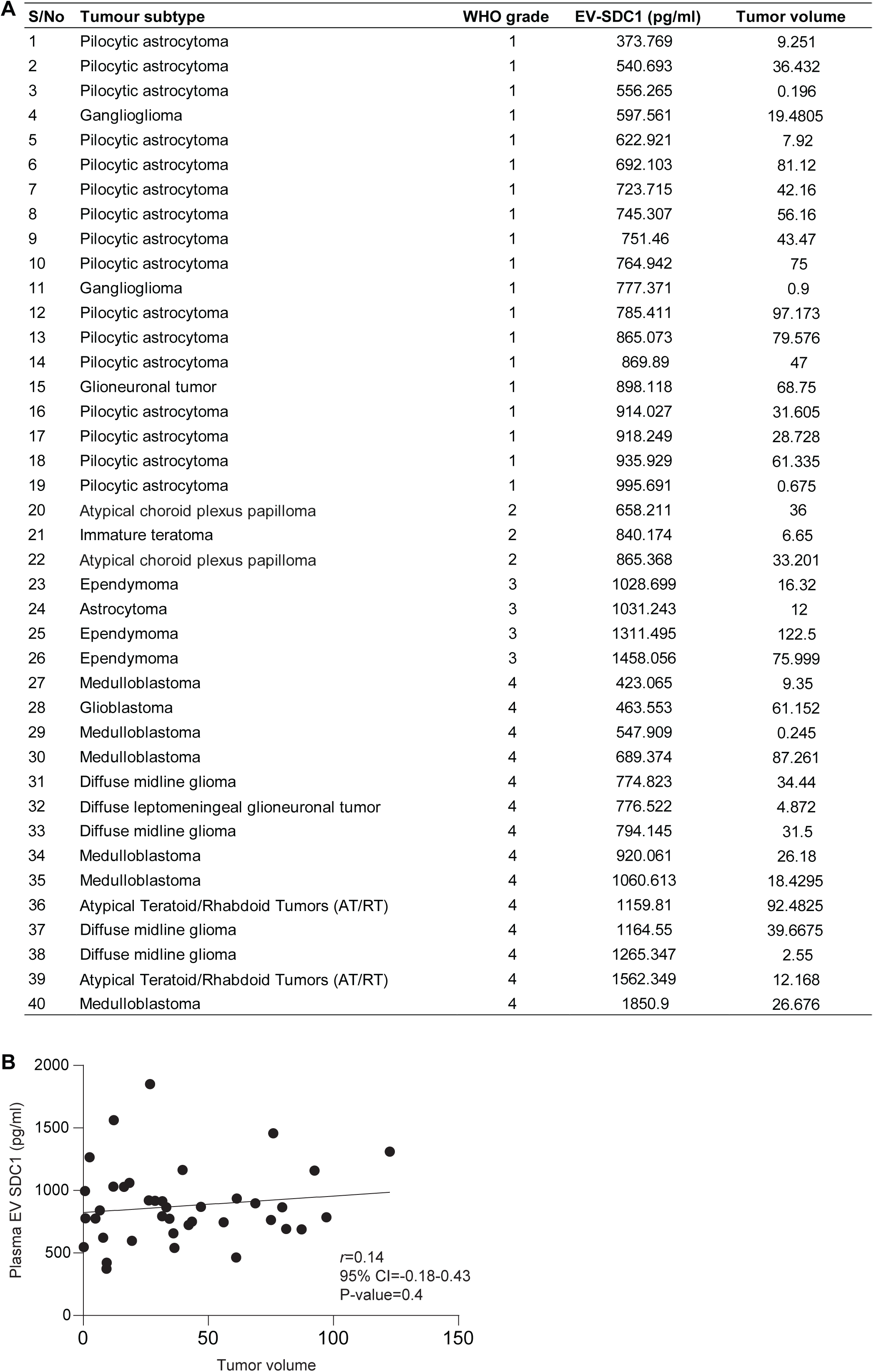
Measurement of patient tumor volume. (A) Table showing tumor volume of different patient tumor subtypes measured from MRI. (B) Correlation plot of extracellular vesicle-syndecan 1 (EV-SDC1) vs tumor volume.

